# Quantifying impacts of the COVID-19 pandemic through life expectancy losses: a population-level study of 29 countries

**DOI:** 10.1101/2021.03.02.21252772

**Authors:** José Manuel Aburto, Jonas Schöley, Ilya Kashnitsky, Luyin Zhang, Charles Rahal, Trifon I. Missov, Melinda C. Mills, Jennifer B. Dowd, Ridhi Kashyap

**Author notes:** For correspondence: José Manuel Aburto, Jonas Schöley, and Ridhi Kashyap. The replication files for this paper including customised functions in the statistics environment R are available on Zenodo, a general-purpose open-access repository developed under the European OpenAIRE program and operated by CERN (10.5281/zenodo.4556982). We would like to thank Alyson van Raalte, Jim Oeppen, Christiaan Monden and John Ermisch for their insightful comments on the manuscript. We also thank Ainhoa Alustiza for her timely response on issues related to the STMF dataset. Funding is gratefully acknowledged from the British Academy’s Newton International Fellowship grant NIFBA19/190679 (JMA, RK), a Rockwool Foundation’s Excess Deaths grant (JMA, JS, TIM, IK), a Leverhulme Trust Large Centre Grant (JMA, LZ, RK, CR, JBD and MCM), Nuffield College (JM, RK, CR, JBD, MCM), John Fell Fund grant 0009182 (JMA, RK, CR, JBD, MCM) and European Research Council grant 835079 (MCM).

## Abstract

Variations in the age patterns and magnitudes of excess deaths, as well as differences in population sizes and age structures make cross-national comparisons of the cumulative mortality impacts of the COVID-19 pandemic challenging. Life expectancy is a widely-used indicator that provides a clear and cross-nationally comparable picture of the population-level impacts of the pandemic on mortality. Life tables by sex were calculated for 29 countries, including most European countries, Chile, and the USA for 2015-2020. Life expectancy at birth and at age 60 for 2020 were contextualised against recent trends between 2015-19. Using decomposition techniques, we examined which specific age groups contributed to reductions in life expectancy in 2020 and to what extent reductions were attributable to official COVID-19 deaths. Life expectancy at birth declined from 2019 to 2020 in 27 out of 29 countries. Males in the USA and Lithuania experienced the largest losses in life expectancy at birth during 2020 (2.2 and 1.7 years respectively), but reductions of more than an entire year were documented in eleven countries for males, and eight among females. Reductions were mostly attributable to increased mortality above age 60 and to official COVID-19 deaths. The COVID-19 pandemic triggered significant mortality increases in 2020 of a magnitude not witnessed since WW-II in Western Europe or the breakup of the Soviet Union in Eastern Europe. Females from 15 countries and males from 10 ended up with lower life expectancy at birth in 2020 than in 2015.

## INTRODUCTION

More than 1.8 million lives are estimated to have been lost due to COVID-19 around the world in 2020 (Dong et al., 2020; World Health Organization, 2021). This estimate — while staggering -– masks the uneven impact of the pandemic across different countries and demographic characteristics like age and sex (Dowd et al., 2020), as well as its impact on population health, years of life lost (Pifarré i Arolas et al., 2021), and longevity (Aburto et al., 2021). Moreover, variations in testing capacity coupled with definitional inconsistencies in counting COVID-19 deaths make the true global toll of COVID-19 infections difficult to estimate with accuracy (Karlinsky and Kobak, 2021). To address these measurement challenges, significant efforts have been directed at the harmonisation and analysis of all-cause mortality data. A widely-used approach to quantify the burden of the pandemic using all-cause mortality is through the analysis of excess mortality, defined as the number of deaths observed during the pandemic above a baseline of recent trends (Aburto et al., 2021). Here we go beyond excess deaths and country-specific analyses and focus on the pressing issue of revealing the impacts of the pandemic on life expectancy in a cross-national perspective.

Life expectancy at birth is the most widely-used metric of population health and longevity. It refers to the average number of years a synthetic cohort of newborns would live if they were to experience the death rates observed in a given period throughout their lifespan. This indicator is thus often referred to as ‘period life expectancy’, as it simulates and summarizes the implications of a mortality profile from a calendar year. While the indicator does not describe a cohort’s actual life course (Goldstein and Lee, 2020) and should not be interpreted as a projection or forecast of any individual’s lifespan (Luy et al., 2020; Andrasfay and Goldman, 2021) it provides a timely description of current mortality patterns. The key advantage of period life expectancy arises from the fact that it is age-standardized, making it the preferred indicator for comparisons across countries with populations of different sizes and age structures, and over time(Andrasfay and Goldman, 2021). Life expectancy can also be calculated as conditional on surviving to a given age, e.g. 60, where it refers to remaining life expectancy from age 60. The study of life expectancy in the context of the COVID-19 pandemic matters because it enables us to compare the cumulative impacts of the pandemic against past mortality shocks and recent trends across different countries using a standardized indicator that is routinely monitored to capture differences in mortality.

Prior to the pandemic, life expectancy at birth typically increased almost monotonically in most countries over the 20th and into the 21st century (United Nations, 2021). In recent decades, improvements in life expectancy among high income countries were predominantly driven by gains made at older ages (Aburto et al., 2020), although significant cross-country heterogeneity persists. This heterogeneity has become more prominent since 2010. While some countries in Eastern Europe and the Baltics experienced significant gains in life expectancy in the past decade (Aburto and van Raalte, 2018), others witnessed noticeable slowdowns in the pace of improvements, and in some cases, stalls or even temporary reversal (Ho and Hendi, 2018). For example, life expectancy in the USA (Mehta et al., 2020), England and Wales, and Scotland saw only limited gains in the last decade (Leon et al., 2019; Fenton et al., 2019). These atypical trends have been linked to slower improvements in old-age mortality, and increases in working-age death rates (Leon et al., 2019).

In a context where trajectories of life expectancy progress became more varied, the COVID-19 pandemic triggered a global mortality crisis posing additional challenges on population health. Death rates from COVID-19 tend to be higher among males than females, with higher case-fatality rates among older age groups (Levin et al., 2020; Ahrenfeldt et al., 2021), precisely those that have accounted for mortality improvements in recent years. The pandemic also indirectly affected mortality from other causes of death. Emerging evidence has highlighted the impacts of delayed treatments or avoidance of care-seeking for cancers or cardiovascular diseases (Hanna et al., 2020; Wu et al., 2021) resulting in increased mortality from these conditions, while lockdowns may have reduced the number of deaths due to accidents (Calderon-Anyosa and Kaufman, 2021).

This study is the first to use an unprecedented collection of demographic data from 29 countries, representing most of Europe, Chile and the USA, to examine the impacts of the pandemic on life expectancy in 2020, contextualised against trends in 2015-2019. To enable reliable cross-national comparisons of life expectancy over the period 2015-2020, we harmonised death counts and population estimates from multiple sources, leveraging major ongoing efforts collecting data on all-cause mortality (Max Planck Institute for Demographic Research, 2021) and official COVID-19 deaths (Riffe et al., 2021). An interactive dashboard that accompanies our manuscript is available at https://covid19.demographicscience.ox.ac.uk/lifeexpectancy. Only countries with high quality age-disaggregated all-cause mortality data were included to estimate life tables (see Methods). We focus on the pressing questions of how much life expectancy changed in 2020 relative to the period 2015-2019, and whether the impact was different for males and females. Leveraging demographic decomposition methods, we examine which age-groups contributed to changes in life expectancy in 2020, and to what extent observed reductions in life expectancy were attributable to officially reported COVID-19 deaths.

## METHODS

### Data

All-cause death counts were retrieved from the Short Term Mortality Fluctuations (STMF) data series within the Human Mortality Database (HMD) (Max Planck Institute for Demographic Research, 2021). Additional all-cause mortality data were retrieved for the USA from the CDC (Centers for Disease Control and Prevention, 2021) and England & Wales from the ONS (Office for National Statistics, 2021) to supplement the STMF data. STMF provides high-quality weekly death counts for 38 countries (at the time of writing) in both a harmonised and original format, but the completeness of these data varies by country. We processed the original input files and selected 29 countries for further analysis. We selected only those countries for which sex-specific death counts across at least 10 age groups were available for every week in 2020 (see Table S1). Death counts for the USA and England & Wales across the years 2015 to 2019 were only reported in extremely coarse age groups by the STMF, but were available in fine grouping for 2020. For both countries we therefore utilised age specific annual death counts as reported by the CDC (Centers for Disease Control and Prevention, 2021) and the ONS (Office for National Statistics, 2021) for the years prior to 2020. All-cause mortality data were supplemented with COVID-19 deaths from the COVerAGEDB database for a subset of 18 countries (by sex) registered continuously since March 2020 until the end of the year (Riffe et al., 2021). COVerAGE-DB provides information about COVID-19 confirmed cases and deaths as reported by statistical agencies for over 100 countries, in a standardised format with harmonised age groups. Population estimates were retrieved from the United Nations for the years 2015-2020; and for the year 2020 we used age-specific mid-year population projections from World Population Prospects 2019 (United Nations, 2021). For England & Wales, Northern Ireland and Scotland, mid-year population estimates from HMD for the year 2018 were projected for 2019 and 2020 using stable population assumptions (see Supplementary Figure S1). We opted against official projections because the WPP does not disaggregate the UK into its constituent countries, and the oldest age group in ONS projections is 90+.

### Mortality estimation

All-cause mortality data are available in irregular age groups for different countries. We harmonised data from STMF, CDC, ONS and COVerAGE-DB using a penalized composite link model (PCLM) which estimated death counts in single years of age from 0 to 110 from the grouped data (Rizzi et al., 2015; Pascariu et al.). The PCLM model was fitted independently to each country, sex, and year combination, and the smoothing parameters were estimated via a Bayesian Information Criteria based grid search. The data points are death counts by age group within each year, country and sex (at least 10 data points for each population). Prior to ungrouping, we summed the weekly death counts from the STMF data into annual death counts, also taking into account deaths reported in unknown weeks. In cases where the age grouping changed from one week to the next, we first summed all the deaths within a stratum and year belonging to the same age grouping scheme, applied PCLM ungrouping separately for each scheme and then summed the ungrouped deaths into annual counts by single years of age.

Person-years of exposure were approximated by estimated or projected mid-year population counts and used in the denominator of the age-specific death rates. Exposure estimates were adjusted for leap-weeks as most countries in the STMF data report deaths using the ISO week date calendar (International Organization for Standardization, 2021). The length of a year in the ISO week calendar is either 371 days in a leap-week year such as 2020, or 364 days in a regular year such as 2019. Thus, the longer reporting interval for leap-week years would, other things equal, increase observed death counts by a factor of 371/364=1.9% over regular years. To counter this spurious effect, we adjusted the exposures by the factor 371/364 in ISO week date leap years. This adjustment has been made for all countries except for annual deaths prior to 2020 for the USA and England & Wales, which are reported over a Gregorian year by the CDC and ONS sources.

### Life table construction

Life tables for all 29 countries by sex for the period 2015-2020 were constructed following a piece-wise constant hazard model using all-cause mortality by single age with the last age interval grouping deaths at ages 85+, consistent with standard demographic techniques (Preston et al., 2001). Our choice of grouping deaths above age 85+ is common practice in demographic research, and it ensures comparability across countries, especially because some countries used this last age group to report deaths for 2020. The choice of the last open-ended interval may impact estimates of life expectancy depending on the proportion of the population surviving to this last interval (Missov et al., 2016). To evaluate its impact, we further calculated life tables by grouping death counts estimated from the PCLM model above age 100+ for countries where more granular age categorisation was available (see Supplementary Figure S2). Smaller populations were more sensitive to the choice of the last open-ended group. However, trends in life expectancy remained similar. From these life tables, life expectancy at birth and life expectancy conditional on surviving to age 60 for males and females were extracted for each country by sex. We performed several sensitivity checks to ensure the robustness of our estimates including comparisons with alternative sources such as the United Nations and from the Human Mortality Database for the period from 2015 to 2019, or their most recent year available (see Supplementary Figure S2). Additionally, we compared full year death counts in our estimates with those reported in the HMD for previous years (Supplementary Figure S3). Most countries did not show deviations across sources, except for Austria, where the STMF underestimates death counts varying from 2% and 4% for females and males, respectively. We further calculated life expectancy using the death counts from both sources and WPP population estimates as exposures and found that the effect on life expectancy is negligible (less than 1%). For the USA, we also compared our life expectancy estimates with those based on provisional data up to June 2020 (Arias et al., 2021). Our results show a sharper decrease of life expectancy than those based on midyear estimates, in line with including the remaining months of 2020 that also saw persistent levels of excess mortality.

### Confidence intervals

We performed Poisson sampling of age-specific death counts to generate confidence intervals around the change in life expectancy from 2019 to 2020 (Silcocks et al., 2001). For each country, sex, year, and age, we sampled 500 death counts from a Poisson distribution with a rate parameter equal to the expected death count predicted by the PCLM method. We calculated a complete life table for each of these 500 replications and derived the annual changes in life expectancy. The sampling distribution of these life expectancy changes was summarised by the 2.5% and 97.5% quantiles, forming a 95% confidence interval.

### Decomposition of life expectancy by age and cause of death

To disentangle age-specific effects and to quantify the impact of COVID-19 deaths on changes in life expectancy, we used the linear integral decomposition method (Horiuchi et al., 2008), a state-of-the-art method that allows us to decompose the difference of two values of life expectancy by age and cause of death, which has been implemented previously for this type of analysis (Aburto and van Raalte, 2018). This methodology assumes that causes of death are exhaustive and independent. This assumption may not be realistic in the context of COVID-19, as the pandemic may have indirectly affected other causes of death. However, previous evidence suggests that the net effect of interactions between causes of death are negligible in decomposition analysis (Beltrán-Sánchez et al., 2008). Moreover, our results are likely to be unaffected by this assumption if COVID-19 deaths are considered ‘excess’. In addition, in most countries the contribution of official COVID-19 deaths to changes in life expectancy can be interpreted as a lower bound due to limited testing and potential mis-classification of COVID-19 deaths. An alternative to decomposition methods is to quantify ‘Years of Life Lost’ defined as the difference between an individual’s age at death from COVID-19 and their remaining life expectancy at that age had the pandemic not occurred (Pifarré i Arolas et al., 2021). However, this measure of life lost is affected by population age structure and therefore comparisons between countries and over time are not precise. We decompose yearly changes in life expectancy by age and official COVID-19 deaths versus other remaining causes of death for each country by sex. This methodology has the advantage that the resulting estimates are additive and the total change in a given period – such as 2015-2019 – is the sum of yearly estimates from 2015 to 2019. As part of our sensitivity checks, we replicated our results with the step-wise decomposition method (Missov et al., 2016) and quantified the impact of COVID-19 deaths on life expectancy by calculating cause-deleted life tables. (Preston et al., 2001) Results from these analyses with respect to those based on the linear integral model were consistent.

## RESULTS

In our dataset, life expectancy at birth among females in 2019 ranged from 78.6 years in Bulgaria to 86.5 in Spain (Figure 1). Among males, it ranged from 71.4 years in Lithuania to 82.2 in Switzerland. At age 60, countries in Eastern Europe and Scotland exhibited the lowest remaining life expectancy, while older females in France and Spain enjoyed the highest. Female life expectancy was larger than males’ in every country.

**Figure 1:**
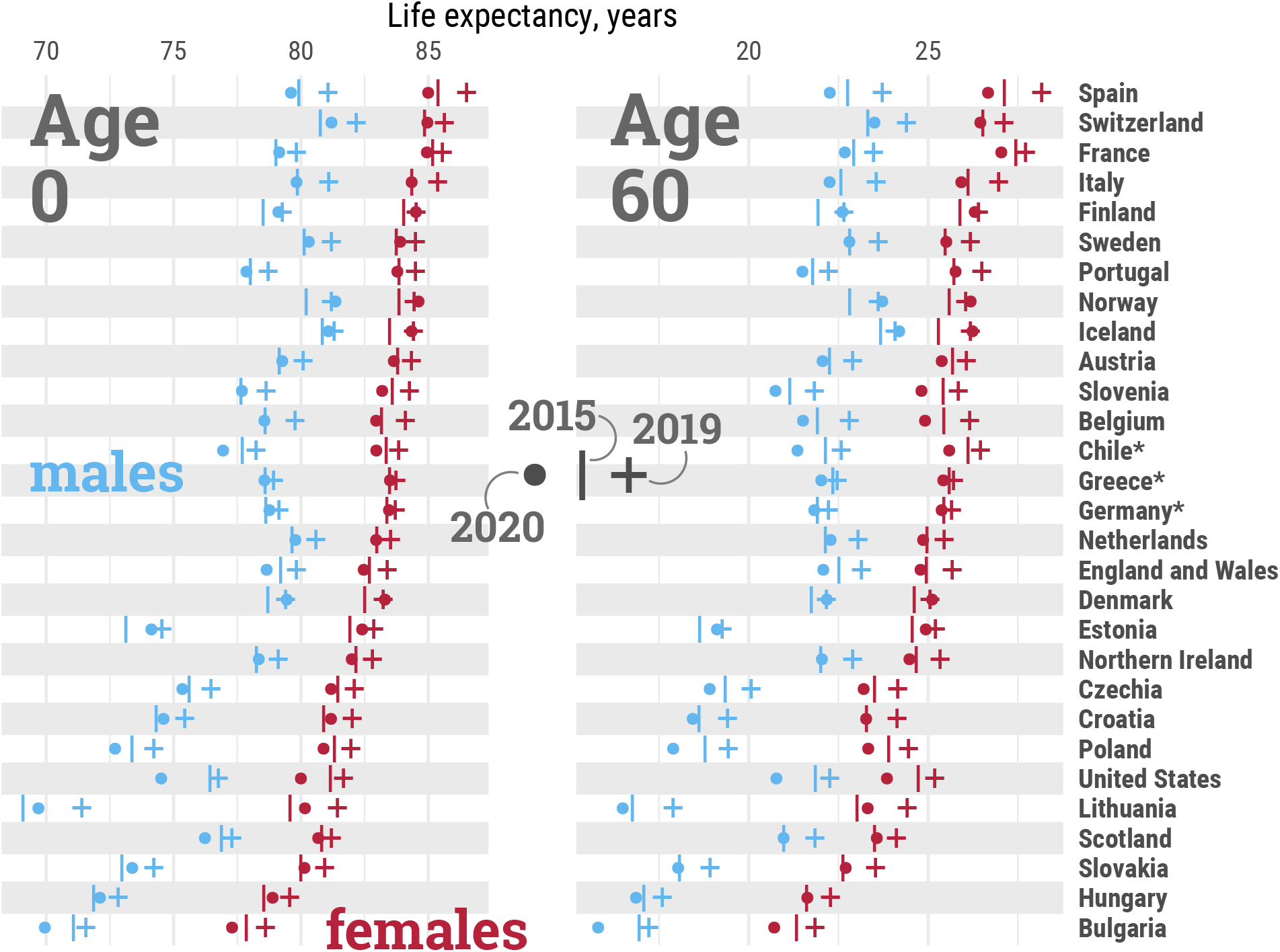
Life expectancy at birth (age 0, left panel) and at age 60 (right panel) by country and sex, in 2015, 2019, and 2020. Note: Estimates for females (red), males (blue), 2015 (|), 2019 (+), 2020 (•) Countries are sorted from highest to lowest levels of female life expectancy at birth in 2019. ^*\**^Estimates for Chile, Greece, and Germany were available from 2016. All data points are provided in a table in Supplementary file 2. An interactive version of this visualisation is available at https://covid19.demographicscience.ox.ac.uk/lifeexpectancy.

From 2015 to 2019, all countries experienced increases in life expectancy at birth, albeit with varying magnitudes (Figure 2). Among females, gains ranged from approximately a month per year in Greece, France, and Scotland, to more than 3 months in Spain, Hungary, and Lithuania. Among males, the lowest gains in life expectancy at birth were seen in the USA, Scotland, and Iceland (up to approximately a month per year), while Lithuanian males benefited from more than five months per year of additional life expectancy. Similar trends across countries were observed for life expectancy at age 60, emphasizing the importance of improvements in older age survival.

**Figure 2:**
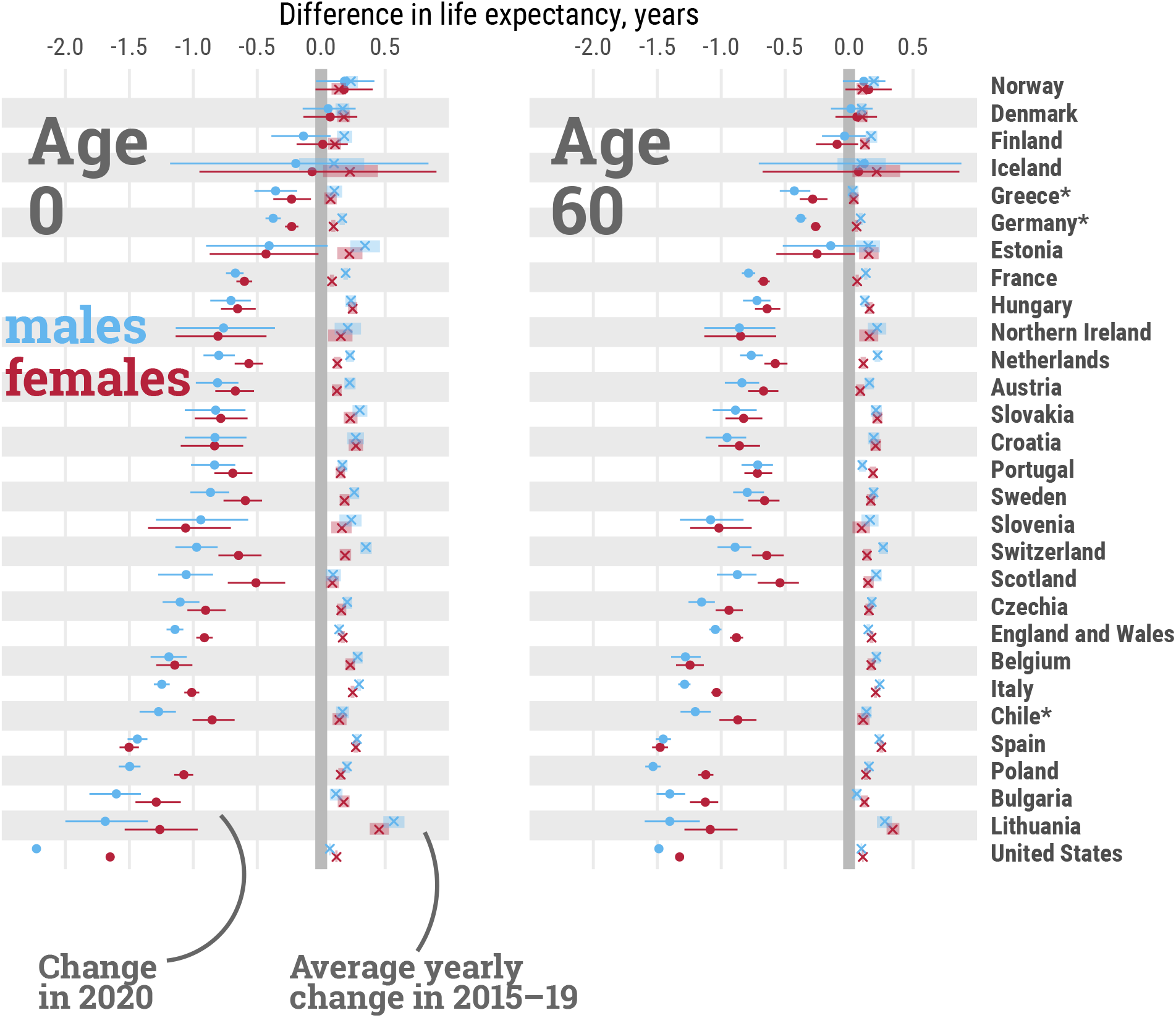
Average per-year change in life expectancy at birth (age 0) and age 60, by country and sex, from 2015 to 2019, and total change from 2019 to 2020. Note: Estimates for females (red), males (blue), average per-year changes from 2015 to 2019 are depicted by the symbol (*x*), dots depict the total change from 2019 to 2020. Lines represent 95% confidence intervals. Countries are sorted from smallest to largest losses between 2019 to 2020 in male life expectancy at birth. ^*\**^Estimates for Chile, Greece, and Germany were available from 2016. All data points are provided in a table in Supplementary file 2. An interactive version of this visualisation is available at https://covid19.demographicscience.ox.ac.uk/lifeexpectancy.

In contrast, life expectancy declined in all countries for both sexes from 2019 to 2020, except for females in Finland and both sexes in Denmark and Norway (Figure 2). With the exception of Spain, Slovenia, Estonia, and Northern Ireland, life expectancy losses were larger for males in 2020. The magnitude of these declines offset most gains in life expectancy in the five years prior to the pandemic. Out of 29 countries, females from 15, and males from 10 countries ended up with lower life expectancy at birth in 2020 than in 2015, which was already an exceptionally bad year for life expectancy due to an irregularly strong flu season (Ho and Hendi, 2018). Our results show that from 2019 to 2020, females in 8 countries and males in 11 lost more than one year of life expectancy at birth. As Figure 3 shows, the magnitudes of losses observed in 2020 have not been witnessed since World War II in many countries of Western Europe such as Spain, England and Wales, Italy, Belgium, France, Netherlands, Sweden, Switzerland and Portugal with data available for the full 20th century (see also Supplementary Figure S4 for females). With the exception of Lithuania and Hungary, the losses observed in 2020 in Eastern European countries exceeded those observed after the dissolution of the Eastern Bloc. The biggest losses of 1.5 years or more of life expectancy at birth were documented among males in the USA, Lithuania, Bulgaria, and Poland, and females in the USA and Spain. At age 60, the three countries with the largest losses in male remaining life expectancy were Poland, USA, and Spain, each losing more than 1.4 years. Remaining life expectancy for females in Spain, USA and Belgium declined by more than 1.2 years. Note that annual changes in life expectancy are subject to random fluctuations that are larger for smaller populations. Thus, the ranking of the countries is not only affected by the pandemic induced mortality shock, but also by random chance, as indicated in Figure 2 via 95% confidence intervals around the estimated drop in life expectancy.

**Figure 3:**
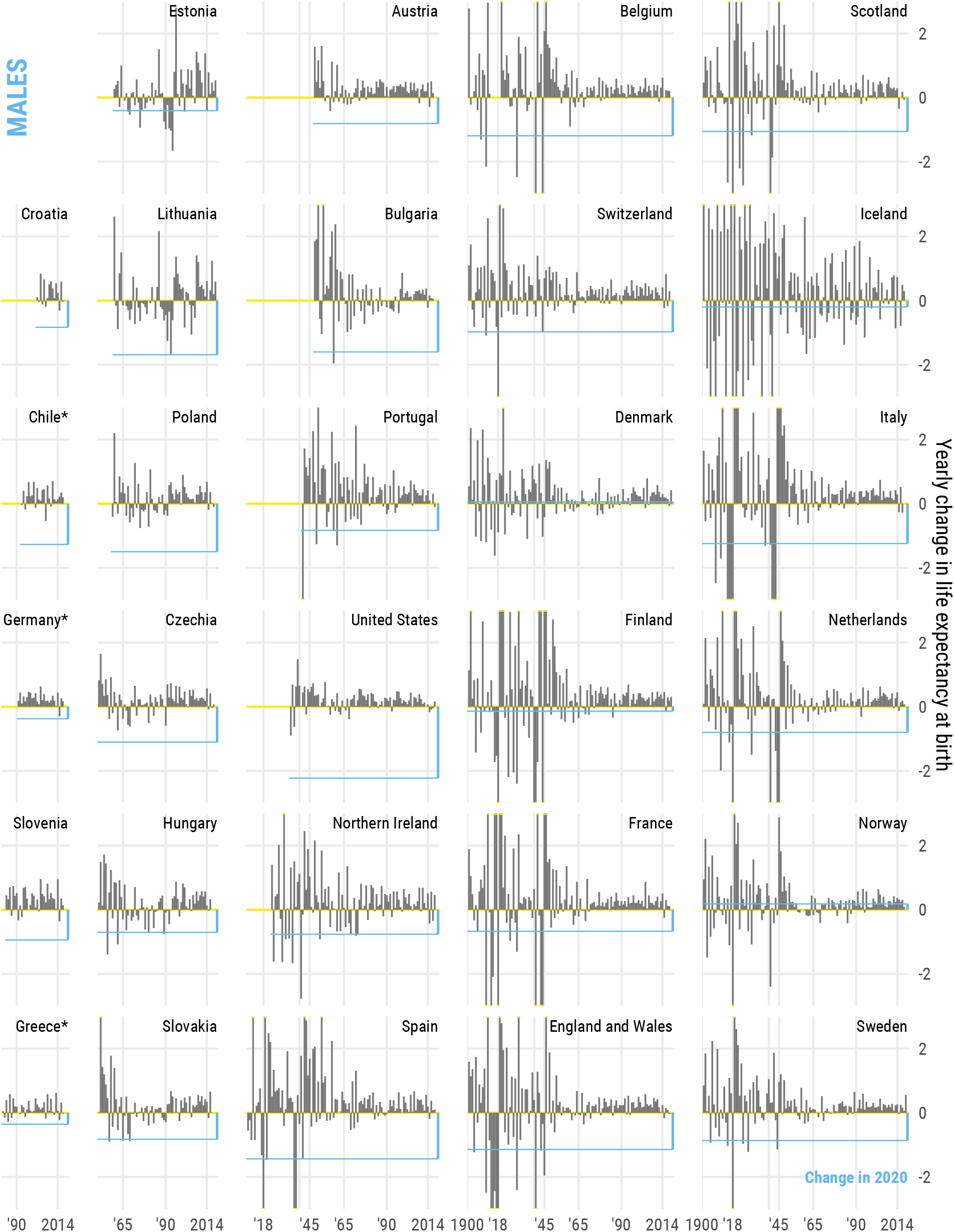
Annual change in male life expectancy at birth since 1900 or earliest year available in the Human Mortality Database for males. The straight blue line indicates the change from 2019 to 2020, with the bars representing year-on-year change.

Mortality reductions are translated into gains in life expectancy at birth and can be attributed to specific age groups. Bars shown in Figure 4 indicate the contributions, in years, to changes in life expectancy in 2015-2019 and 2019-2020 by broad age groups and sex (finer age grouping in Supplementary Figure S5). Between 2015-2019, mortality improvements above age 60 consistently contributed to increasing life expectancy across countries. In several countries, life expectancy gains above age 80 surpassed those in the age group 60-79. For females in particular, mortality improvements in the 80+ age group contributed most to improvements in life expectancy between 2015-19, whereas for men broadly mortality improvements in the 60–79 year age group were greater. Improvements below age 60 contributed less, but progress was still observed in many countries, especially in Eastern Europe, except for the USA and Scotland where life expectancy decreased due to worsening mortality at these ages. The COVID-19 pandemic led to sharp declines in life expectancy (see Figures 2-3), predominantly due to elevated mortality in the older age groups (see Figure 4). Increased death rates above age 60 contributed the most to life expectancy declines across all countries between 2019 and 2020. Increased death rates at ages 80+ contributed most to life expectancy losses among females. Among males, elevated death rates at ages 60-79 contributed most to life expectancy losses across many countries, but the impact of midlife mortality below age 60 was noticeable too, especially those suffering the biggest losses in life expectancy such as the US, Lithuania, and Bulgaria. Notably increased midlife mortality (0-59 years) was the largest contributor to life expectancy losses between 2019 and 2020 in the USA among males.

**Figure 4:**
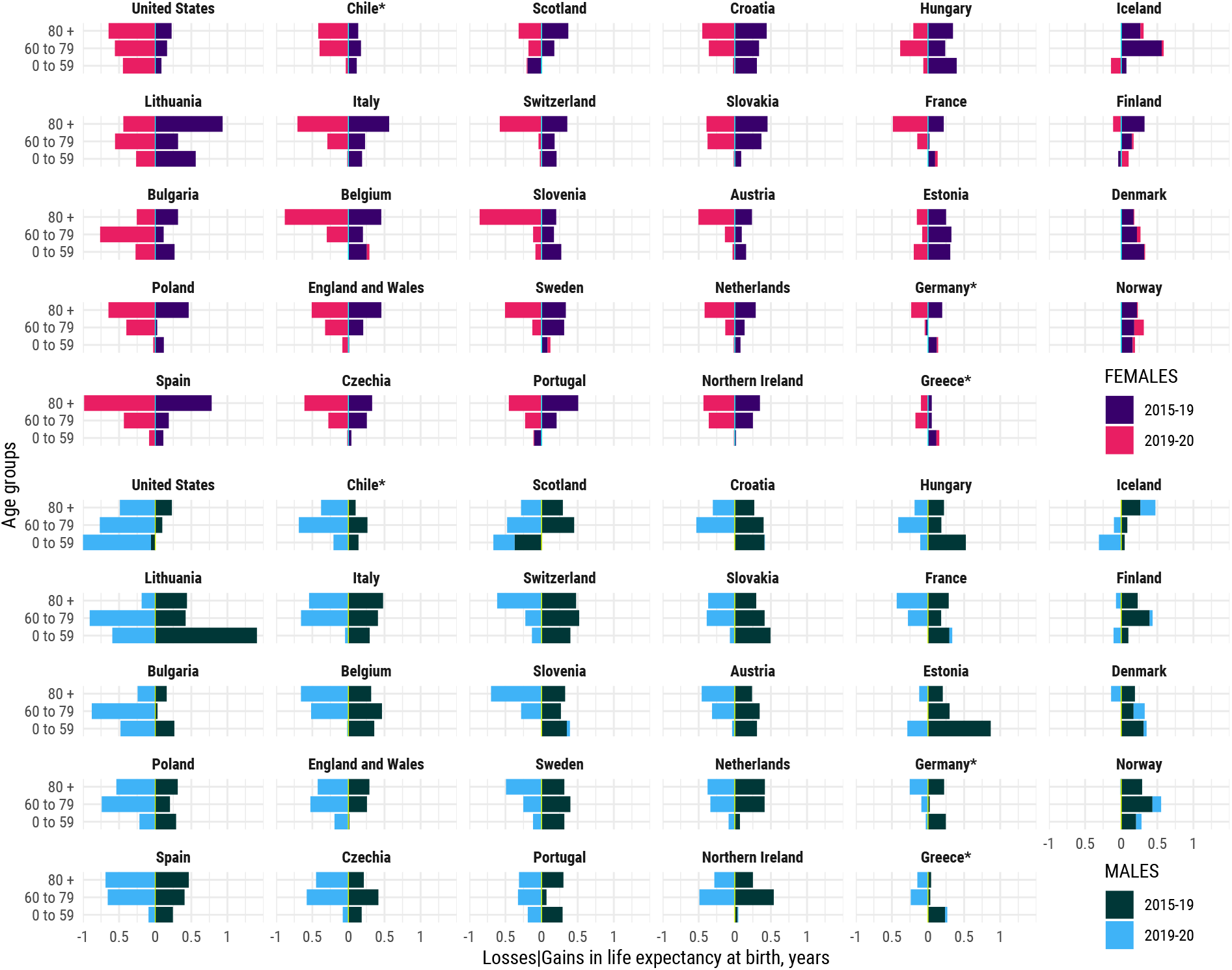
Contributions (in years) to changes in life expectancy at birth from 2015 to 2019, and from 2019 to 2020, attributable to mortality below age 60, from 60 to 80, and above age 80, by country and sex. Note: Positive values indicate gains in life expectancy, negative values indicate reductions in life expectancy. The sum of contributions over age results in the total change of life expectancy at birth during the specific period. Countries are sorted (column-wise) from largest to smallest losses between 2019 to 2020 in male life expectancy at birth.*Estimates for Chile, Greece, and Germany were available from 2016. All data points are provided in a table in Supplementary file 3.

We were able to quantify the contribution of official COVID-19 deaths to life expectancy reductions for 18 countries (Figure 5) for which more reliable age and sex-disaggregated data on COVID-19 deaths were available (see Methods). We estimated that deaths reported as COVID-19 in the COVerAGE database (Riffe et al., 2021) explain most life expectancy losses observed. Both females and males in Chile, England & Wales, Belgium, Spain and Slovenia, and males in Italy experienced losses of more than a whole year of life expectancy at birth attributable to official COVID-19 deaths, explaining most of the overall loss between 2019 and 2020. To put this into perspective, it took on average 5.6 years for these countries to achieve a one-year increase in life expectancy recently: progress wiped out over the course of 2020 by COVID-19.

**Figure 5:**
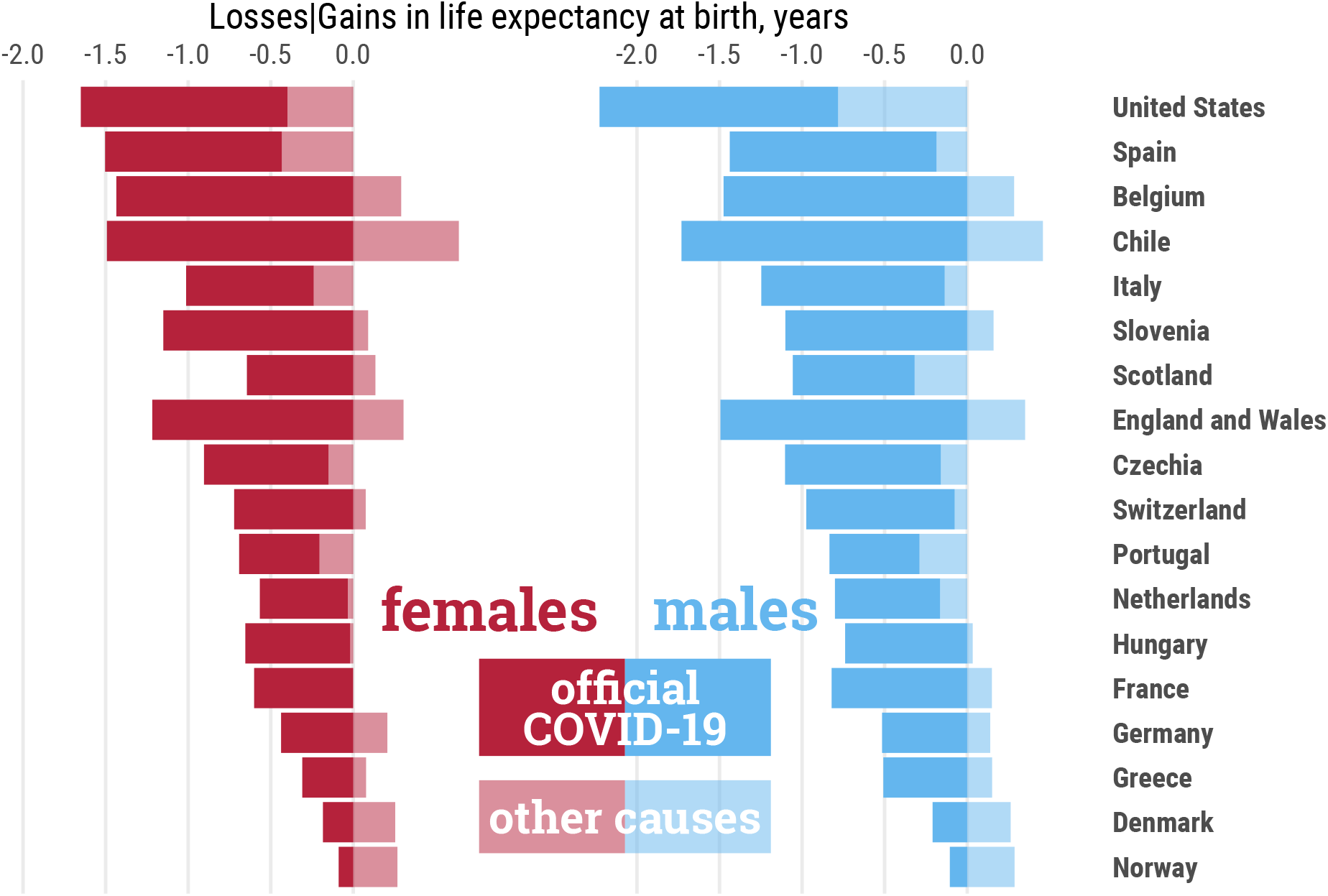
Contributions (in years) to changes in life expectancy at birth from 2019 to 2020 attributable to official COVID-19 deaths and remaining causes of death. Note: Countries are sorted from largest to smallest losses. The sum of both components adds to the total change from 2019 to 2020 in a given country. All data points are provided in a table in Supplementary file 3

In some contexts, such as Denmark and Chile, life expectancy losses due to COVID-19 deaths were larger than total life expectancy losses, as increased mortality due to COVID-19 was offset by mortality reductions among other causes.

### Limitations

While most of the countries included in our analysis have high-quality systems of death registration and usually report timely and complete death counts (Barbieri et al., 2015), there are likely issues related to delays in death registration in 2020. We deal with these potential issues by harmonizing and smoothing death counts with a penalized composite link model which has been shown to provide reliable estimates of mortality for all countries in our study (Rizzi et al., 2015), and by adjusting the person-years exposed. In most STMF countries, deaths that take place in a given week are covered by more than 90% by the following two weeks (STMF metadata). Therefore, we expect the percentage of missing deaths in the last weeks of 2020 to be low. Moreover, according to the STMF metadata, some countries (such as Austria, England and Wales, Germany, and the Netherlands) employ extrapolation procedures to correct for incompleteness to adjust for missing deaths counts in the preceding 1-to-3 weeks. We recognise nonetheless that late and/or under registration may affect our estimates by underestimating losses in 2020. Therefore, our results should be interpreted as estimates of the lower bound of life expectancy reductions in 2020. For official COVID-19 mortality figures, there are likely to be inaccuracies due to differences in classification and testing practices across countries. While we are unable to precisely measure the extent to which these inaccuracies may affect our estimates to make adjustments (i.e., since mis-classification can occur for different reasons), we err on the side of caution and interpret our results as a lower bound of the impact of COVID-19 deaths on changes in life expectancy.

The magnitude of life expectancy losses can also be influenced by migration due to COVID-19, especially in countries where persistent out-migration flows are not tracked accurately (such as via population registers) between censuses. For example, in Eastern European countries with high levels of out-migration in their populations, increased mortality could have occurred because of elevated return migration due to the COVID-19 pandemic. Nevertheless, we anticipate that the magnitude of these effects is likely to be small due to the broadly younger age profiles of migrants and previous evidence that suggests that mortality at younger ages is more likely to be affected by international migration (Rogers and Castro, 1981; Raymer and Wiśniowski, 2018). In 2020, as increased mortality occurred mostly at older ages, the effect of migration on our estimates is likely to be low.

## DISCUSSION AND CONCLUSION

Our analyses of life expectancy show that the pandemic exacted a striking toll on population health in 2020 across most of Europe, the USA and Chile. Only males and females in Denmark and Norway, and females in Finland were successful in avoiding drops in life expectancy in our cross-national comparison of 29 countries. Early non-pharmaceutical interventions coupled with a strong health care system may help explain some of this success (Conyon et al., 2020; Juranek and Zoutman, 2021). In contrast, the USA, followed by Eastern European countries such as Lithuania, Bulgaria, Poland, experienced the largest losses in life expectancy in 2020, with larger losses in most countries for males than females. Among Western European countries, life expectancy losses were also documented. To put this into perspective, 2014-15 strong flu season was regarded as an exceptionally bad year among high income countries, causing drops in life expectancy (Ho and Hendi, 2018). The largest drop was observed in Italy with a loss of around half a year among males and females. Our study reveals that 22 countries included in our study experienced larger losses than half a year in 2020. Females in 8 countries and males in 11 countries experienced losses larger than a year. For Western European countries such as Spain, England and Wales, Italy, Belgium, among others, the last time such large magnitudes of losses were observed in a single year was during WW-II.

While in Europe, increased mortality in the 60+ age groups contributed most to life expectancy losses, the pandemic saw a worsening of working-age mortality (under 60 years) in the USA, and this age group contributed most to life expectancy losses for males. Despite being a younger population, the USA also has higher co-morbidities in these age groups compared with European populations with greater vulnerability to COVID-19 (Avendano and Kawachi, 2014; Lawrence et al., 2018). Other factors, such as those linked to unevenness in healthcare access in the working-age population and structural racism may also help explain increased mortality. Recent research from the USA, for example, shows socially disadvantaged populations such as Blacks and Latinos experienced losses three times higher than those reported here at the national level9. Emerging evidence further indicates that non-COVID-19 excess mortality was concentrated in working ages (Faust et al., 2021; Glei, 2021). While some of this likely reflects mis-classification of COVID-19 deaths due to factors linked to poorer testing and healthcare access, our findings reflect a deepening during the pandemic of the USA mid-life mortality crisis, that has led to the stalling of life expectancy in recent years (Ho and Hendi, 2018; Mehta et al., 2020; Preston and Vierboom, 2021).

Countries in Eastern Europe (Lithuania, Bulgaria and Poland), as well as England and Wales and Scotland, where recent life expectancy stalls due to deteriorating midlife mortality too have been noted, also saw increased mortality in the under-60 age group. In the Eastern European countries, increased mortality in the 60-79 age group contributed most to life expectancy losses in comparison with Western European countries such as, for example, Spain, Italy, Belgium, where increased mortality in 80+ age groups had sizable impacts. These patterns may reflect the continued impacts of the atypical mortality transitions and a delayed cardiovascular revolution in Eastern countries compared with Western Europe (Aburto and van Raalte, 2018), whereby greater co-morbidities and vulnerabilities in population health are present at these ages.

Emerging evidence from low- and middle-income countries (such as Brazil and Mexico) which have been devastated by the pandemic (Karlinsky and Kobak, 2021) suggests that life expectancy losses may be even larger in these populations. Losses in life expectancy are also likely to vary substantially between subgroups within countries. However, a lack of data currently limits direct and more disaggregated comparisons across a wider range of countries, but these are urgently needed to understand the full mortality impacts of the pandemic. This limitation is manifested in our study as we were only able to include two countries outside of Europe with reliable and complete information for 2020, and our analysis focused on national populations. To expand our analyses to a broader set of countries, data on all-cause mortality with detailed disaggregation by age and sex, and for deeper analysis other characteristics such as socioeconomic status and ethnicity are needed to assess the uneven mortality impacts of the pandemic. Nevertheless, we note that in settings with deficient or limited vital registration the true mortality impacts of the pandemic may never fully be known, and at minimum, use of data from such contexts would require further adjustments (e.g. for under-counting, age heaping) before use. While COVID-19 might be seen as a transient shock to life expectancy, the evidence of potential long-term morbidity due to long-COVID and impacts of delayed care for other illnesses (Hanna et al., 2020; Wu et al., 2021) as well as health effects and widening inequalities stemming from the social and economic disruption of the pandemic (Bambra et al., 2020) suggest that the scars of the COVID-19 pandemic on population health may be longer lasting.

## Data Availability

The replication files for this paper including customised functions in the statistics environment R are available on Zenodo, a general-purpose open-access repository developed under the European OpenAIRE program and operated by CERN (10.5281/zenodo.4556982).

https://github.com/OxfordDemSci/ex2020

## Authors contributions

Conceptualization: JMA, JS, RK. Data curation: JS, LZ, JMA, RK, TIM, IK. Formal analysis: JMA, JS, LZ, IK. Methodology: JMA, RK, JS, IK. Software: JS, JMA, LZ, IK. Visualization: IK, JS, CR, JMA, RK, JBD, MCM. Project administration: JMA, RK. Supervision: JMA, RK, JS. Drafting: JMA, RK. Review and editing: LZ, JS, IK, TIM, CR, JBD, MCM.

## Data and materials availability

The replication files for this paper include customised functionality written in the R statistical programming language (R. Core Team, 2013). The code, and all harmonized input and output data pertaining to our analysis, is hosted both on Zenodo (a general-purpose open-access repository developed under the European OpenAIRE program and operated by CERN) at https://zenodo.org/record/4556982, and on GitHub (https://github.com/oxforddemsci/ex2020).

