## Supplementary File 1 for "Quantifying impacts of the COVID-19 pandemic through life expectancy losses: a population-level study of 29 countries"

José Manuel Aburto[‡,§,*], Jonas Schöley[*,†], Ilya Kashnitsky[*], Luyin Zhang[‡,¶]
Charles Rahal[‡,§], Trifon I. Missov[*], Melinda C. Mills[‡,§],
Jennifer B. Dowd[‡,§], Ridhi Kashyap[‡,§,†]

[‡] *Leverhulme Centre for Demographic Science and Department of Sociology, University of Oxford*
[§] *Nuffield College, University of Oxford*
[*] *Interdisciplinary Centre on Population Dynamics, University of Southern Denmark*
[†] *Max Planck Institute for Demographic Research, Rostock*
[¶] *St. Cross College, University of Oxford*

6th August, 2021

| Country/Region | All cause death counts # Groups 2019 | # Groups 2020 | Miss. 2020 | Open-age 2019 | Open-age 2020 | Covid deaths Latest report 2020 | Notes |
|---|---|---|---|---|---|---|---|
| Austria | 19 | 19 | 0 | 90 | 90 | 31/12/2020 | COVID death counts not available by sex |
| *Australia* | 5 | 5 | 0 | 85 | 85 | 31/12/2020 | Coarse ages. |
| **Belgium** | 19 | 19 | 0 | 90 | 90 | 31/12/2020 | |
| Bulgaria | 19 | 19 | 0 | 90 | 90 | / | COVID death counts unavailable. |
| *Canada* | 4 | 4 | 0 | 85 | 85 | 22/09/2022 | Coarse ages. |
| **Switzerland** | 19 | 19 | 0 | 90 | 90 | 31/12/2020 | |
| **Chile** | 20 | 20 | 0 | 95 | 95 | 31/12/2020 | No data for 2015. |
| **Czech Republic** | 19 | 19 | 0 | 90 | 90 | 31/12/2020 | |
| **Germany** | 15 | 15 | 0 | 95 | 95 | 31/12/2020 | No data for 2015. |
| **Denmark** | 21 | 21 | 0 | 100 | 100 | 31/12/2020 | |
| Estonia | 19 | 19 | 0 | 90 | 90 | / | COVID death counts unavailable. |
| **Spain** | 19 | 19 | 0 | 90 | 90 | 31/12/2020 | |
| Finland | 19 | 19 | 0 | 90 | 90 | / | COVID death counts not available by sex. |
| **France** | 21 | 21 | 0 | 95 | 90 | 31/12/2020 | |
| **England & Wales** | 106 | 20 | 0 | 100 | 90 | 25/12/2020 | Pre-2020 data from ONS. |
| Northern Ireland | 19 | 19 | 0 | 90 | 85 | / | Last 2 weeks with 7 age-groups. COVID deaths not available by sex. |
| **Scotland** | 20 | 21 | 0 | 90 | 95 | 31/12/2020 | |
| *Greece* | 19 | 19 | 0 | 90 | 90 | 31/12/2020 | No data 2015. |
| Croatia | 19 | 19 | 0 | 90 | 90 | / | |
| **Hungary** | 19 | 19 | 0 | 90 | 90 | 31/12/2020 | |
| *Israel* | 8 | 8 | 0 | 80 | 80 | 28/12/2020 | Coarse ages. |
| Iceland | 19 | 19 | 0 | 90 | 90 | / | |
| **Italy** | 22 | 19 | 0 | 100 | 90 | 29/12/2020 | Missing weeks. |
| *South Korea* | 5 | 5 | 0 | 85 | 85 | 28/06/2020 | Coarse age grouping. COVID deaths after 2020-06-28 not available by sex. |
| Lithuania | 19 | 19 | 0 | 90 | 90 | / | COVID death counts unavailable. |
| *Luxembourg* | 19 | 19 | 0 | 90 | 90 | / | Low number of younger deaths and small pop sizes produced atypical shape of hazard with PCLM. COVID death counts unavailable. |
| *Latvia* | 19 | 19 | 0 | 90 | 90 | / | Low number of younger deaths and small pop sizes produced atypical shape of hazard with PCLM. COVID death counts unavailable. |
| **Netherlands** | 19 | 19 | 0 | 90 | 90 | 27/12/2020 | |
| **Norway** | 21 | 21 | 0 | 100 | 100 | 30/12/2020 | |
| *New Zealand* | 3 | 3 | 0 | 80 | 80 | / | Coarse age grouping. COVID deaths not available by sex. |
| **Poland** | 19 | 19 | 0 | 90 | 90 | / | COVID deaths not available. |
| **Portugal** | 19 | 19 | 0 | 90 | 90 | 31/12/2020 | |
| *Russia* | 19 | / | 53 | 90 | / | / | No data for 2020. COVID deaths not available. |
| Sweden | 19 | 19 | 0 | 90 | 90 | / | COVID deaths not available by sex. |
| **Slovenia** | 19 | 19 | 0 | 90 | 90 | 31/12/2020 | |
| *Slovakia* | 19 | 19 | 0 | 90 | 90 | / | COVID deaths not available. |
| *Taiwan* | 21 | 21 | 1 | 100 | 100 | 10/08/2020 | Missing weeks for 2020. COVID deaths incomplete. |
| USA | 11 | 11 | 0 | 85 | 85 | 26/12/2020 | Pre-2020 data from CDC, aggregated to 2020 age grouping. |

**Table S1: Data quality of raw all-cause and Covid-19 death data.** Note: Source data retrieved 2021-05-25 from HMD-STMF, CoverAge-DB, UK-ONS, and US-CDC. All cause death counts from STMF unless stated otherwise in the notes. Countries noted in italics were excluded from analysis. Countries in bold are part of COVID-19 analysis. ‘Miss.’ refers to latest reported data in 2020.

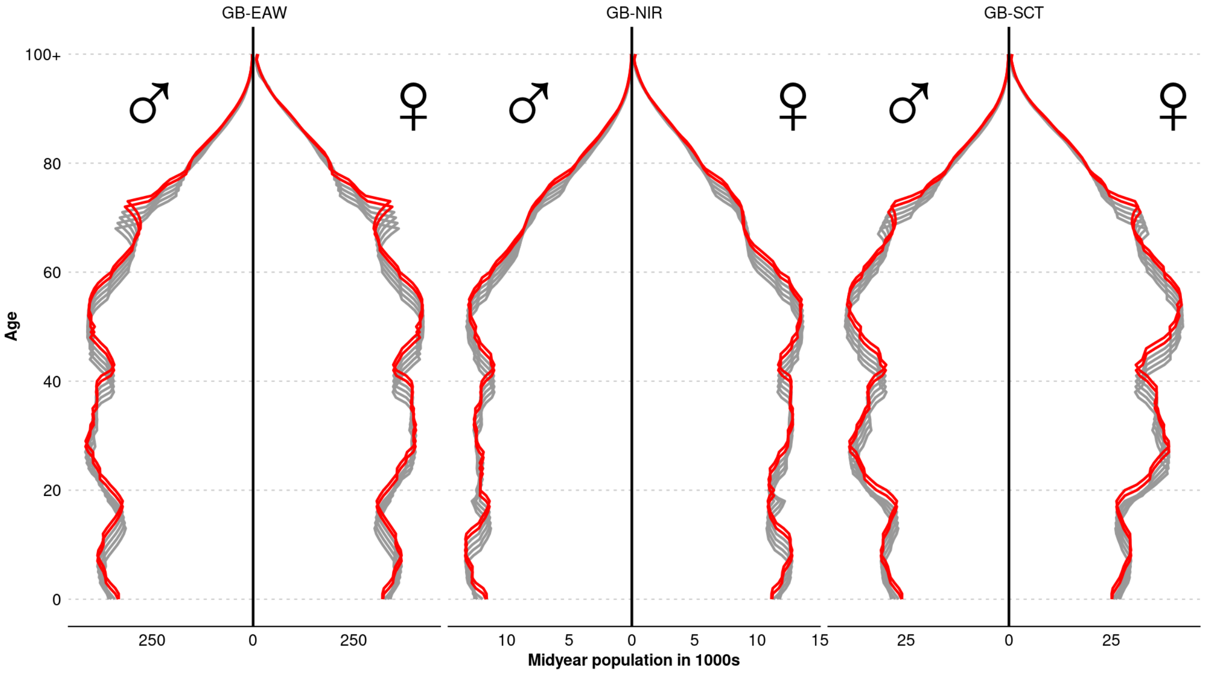


**Figure S1: Midyear population by age 2015-2018 as reported by the Human Mortality Database and own projections for 2019-2020 for England Wales, Scotland and Northern Ireland.**

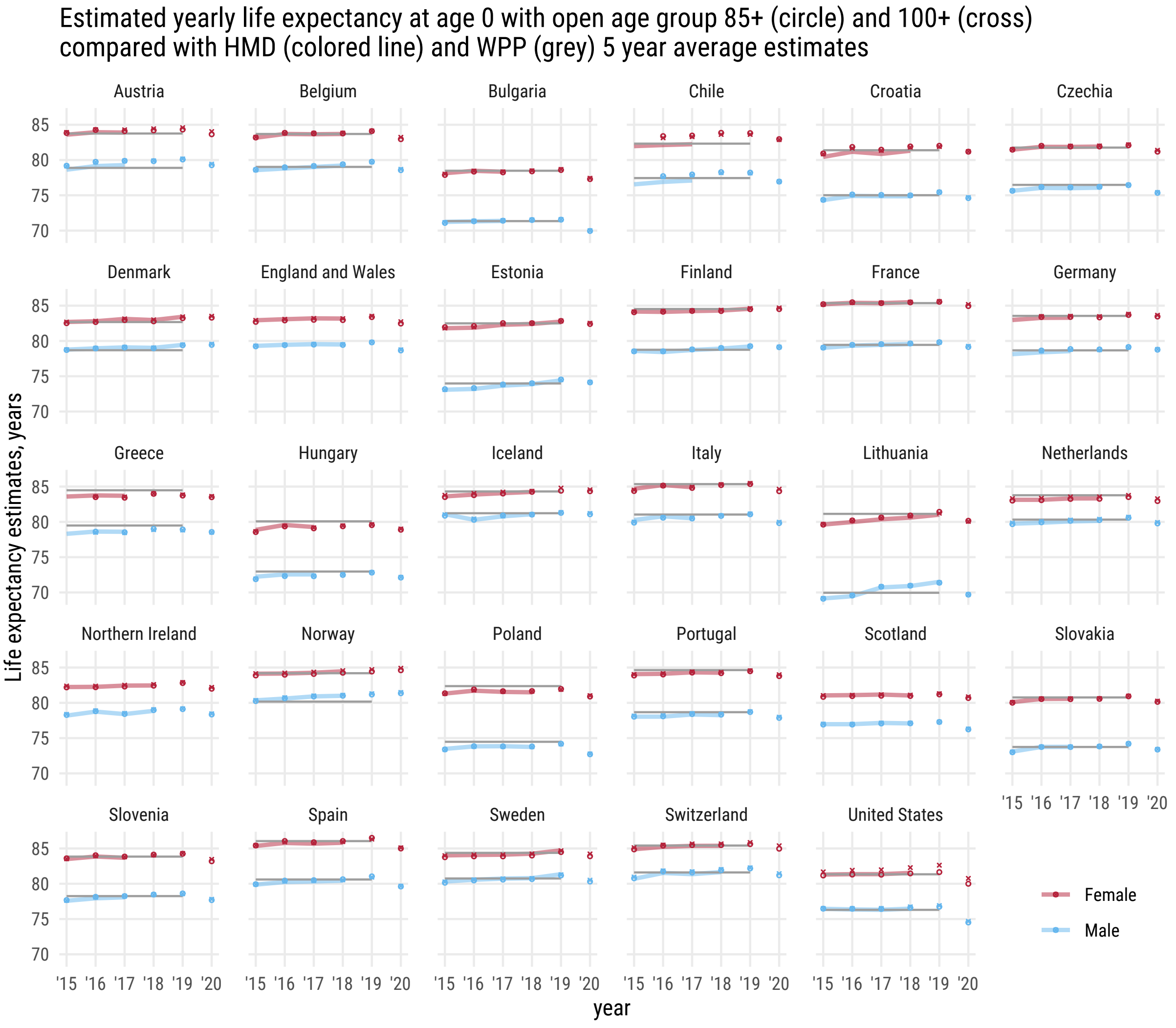


**Figure S2: Comparison of estimates of life expectancy by closing life tables at 85+ and 100+ with those published by the United Nations and the Human Mortality Database in the period 2015-19, or the most updated year available.**

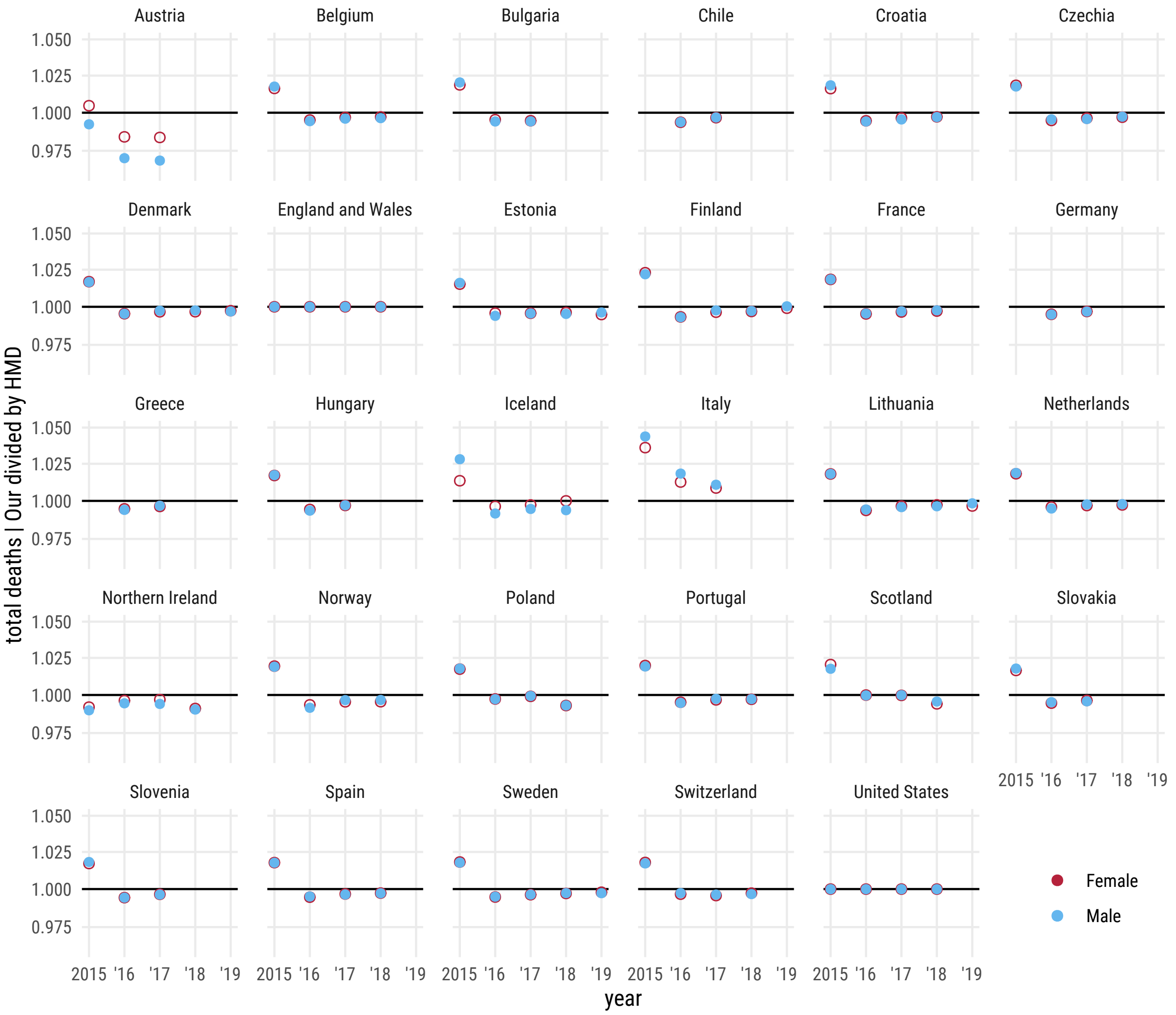


**Figure S3: Comparison of death counts between the Short Term Mortality Fluctuations dataset and the Human Mortality Database.** Note: for discrepancy in 2015 see Methods section on ISO-week formats.

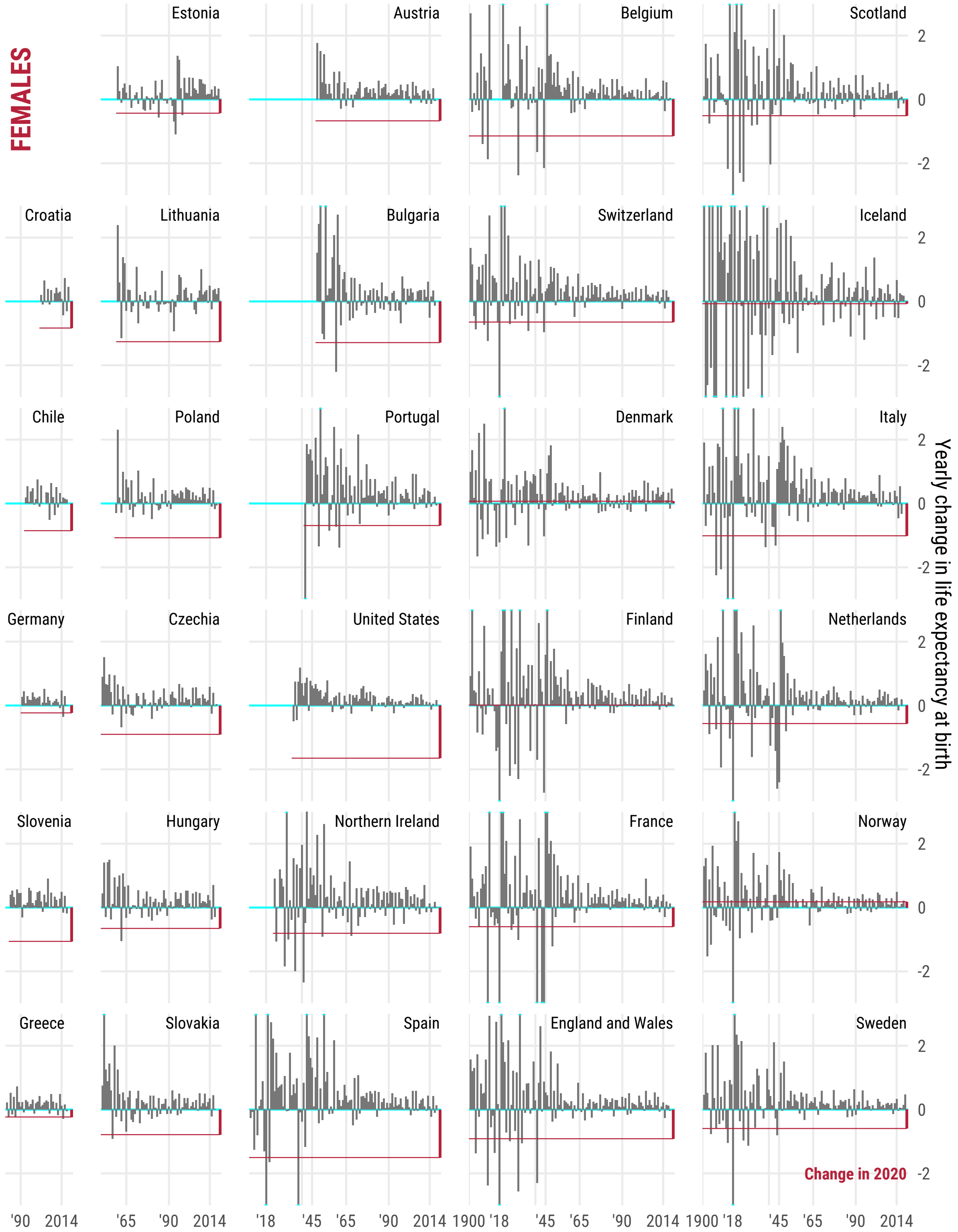


**Figure S4: Annual change in life expectancy at birth since 1900 or earliest year available in the Human Mortality Database for females. The straight red indicates the change from 2019 to 2020, with the bars representing year-on-year change.**

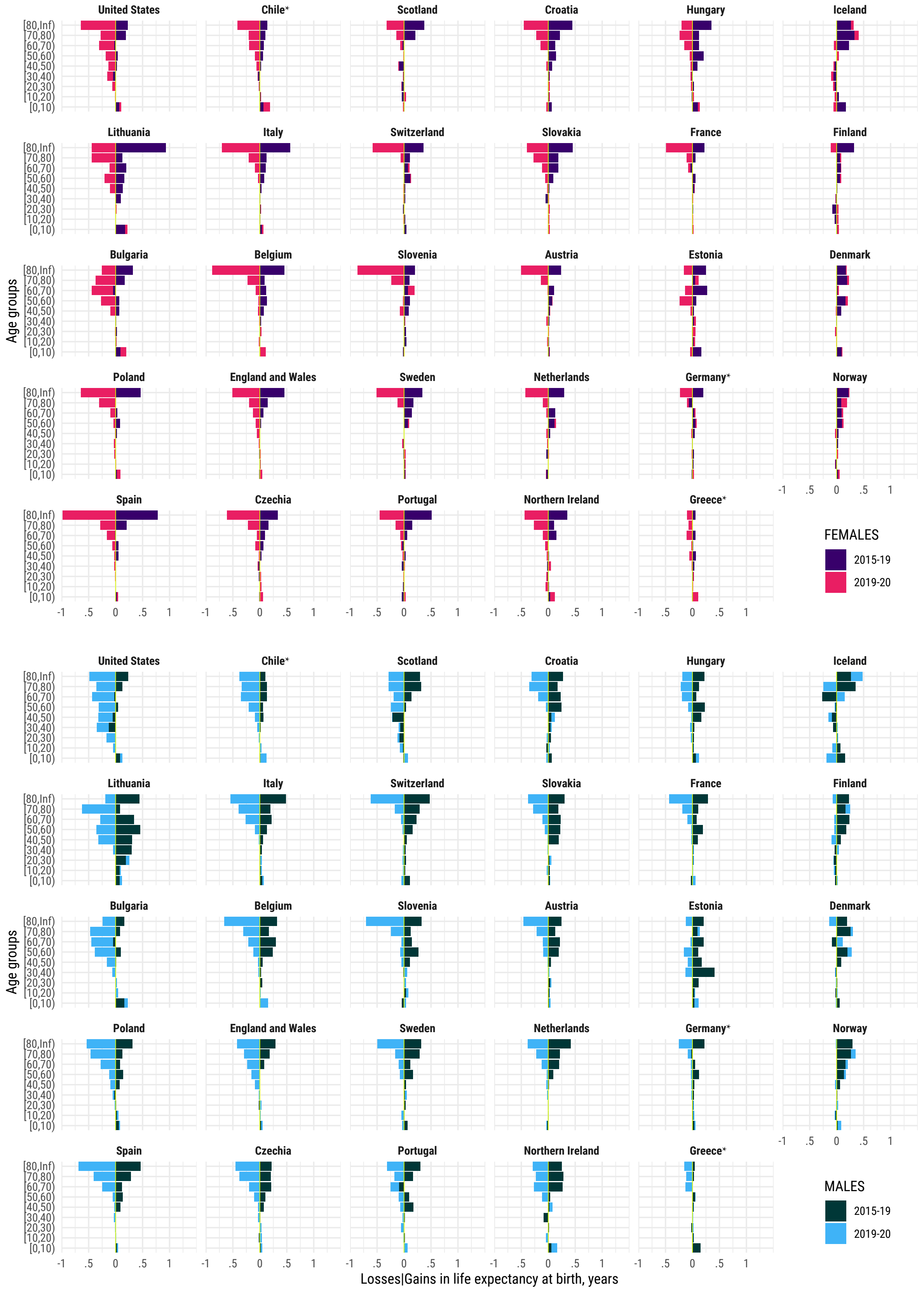


**Figure S5: Contributions (in years) to changes in life expectancy at birth from 2015 to 2019, and from 2019 to 2020, attributable to mortality in different 10-year age groups by country and sex.** Note: Positive values indicate gains in life expectancy, negative values indicate reductions in life expectancy. *Estimates for Chile and Germany were available from 2016.
